# Caregivers’ formal digital proxy roles and engagement in advance care planning: A cross-sectional study

**DOI:** 10.64898/2026.07.08.26357531

**Authors:** Vinh Anh Huynh, Camellia Zakaria, Pavithren VS Pakianathan, Gerald Choon Huat Koh, Pin Sym Foong

## Abstract

Caregivers increasingly act as proxies, managing patients’ digital accounts and making complex end-of-life decisions. Greater dyadic engagement in advance care planning (ACP) improves patient and caregiver outcomes, yet empirical evidence linking formal digital proxy roles to ACP engagement remains limited. The study aims to quantify patterns of ACP engagement, digital proxy roles, and how these caregivers’ behaviors are associated among caregivers in Singapore. We conducted a cross-sectional survey among an online panel of nationally representative adults in Singapore to identify caregivers and assessed their lifetime engagement in formal proxy roles across legal, financial, and medical digital domains, along with ACP proxy behaviors. Formal digital proxies had institutional or joint access to digital financial accounts (for financial digital proxies) or digital patient health/caregiver accounts (for medical digital proxies). ACP engagement was measured using 13 proxy-related behaviors, such as discussing end-of-life care preferences. Multivariable regressions were performed. In total, we identified 276 caregivers, who assisted with instrumental activities daily living to another adult from 311 completed responses. Among caregivers (age 41.0±13.8; 46.2% female), 28.9% were legal proxies and 40.2% were formal digital proxies (31.5% financial; 29.0% medical). Overall engagement was modest (mean 3.97±4.54) despite most reported completing at least one behavior. Compared to non-proxies, medical (AME=3.722, 95%CI: 2.143–5.301) and financial digital proxies (AME=1.515, 95%CI: 0.121–2.910) reported significantly higher ACP engagement while legal proxy status did not. High-stakes discussions on life-sustaining treatment and health-state preferences showed low engagement. Formal digital proxy roles are positively associated with ACP engagement and may provide a strategic entry point for interventions. Persistent deficits in high-stakes ACP highlight limited readiness for complex end-of-life decisions and the need for targeted decision-support tools.

**Author summary:** Informal caregivers often help patients manage digital accounts and may also make important medical care decisions. We examined whether caregivers who hold formal digital proxy roles are more engaged in advance care planning, through behaviors such as discussing proxy roles, patient’s values and preferences, and communicating with doctors. We surveyed 311 adults in Singapore through an online panel representative of the national population and identified 276 caregivers who provided instrumental care for another adult. We asked participants to report their involvement in legal, financial, and medical digital matters, as well as 13 advance care planning behaviors. About 40% of caregivers held formal digital proxy roles, through having access to financial or digital health-related accounts. Overall, advance care planning engagement was modest, with caregivers completing an average of 4 out of 13 behaviors. However, caregivers who served as medical or financial digital proxies reported significantly greater engagement than those without these roles. Legal proxy status alone was not associated with higher engagement. These findings suggest that digital proxy roles may be a practical starting point for helping caregivers move from managing accounts to deeper advance care planning engagement, but caregivers still need more support for difficult end-of-life conversations and decisions.

## Background

Increasing dyadic engagement in advance care planning (ACP) has helped to deliver value-centered end-of-life (EOL) care towards EOL.(1, 2) ACP is described as an ongoing process of planning and preparing for future health and personal care. Recently, ACP research has shifted away from documentation towards a focus on the relational and communicative processes that strengthens shared understanding between patients and caregivers regarding personal beliefs, values and healthcare preferences with the aim to enhance surrogate decision readiness under crisis conditions.(3) This dyadic focus in ACP has been shown to enhance caregiver’s readiness to take on surrogate decision-making roles by helping clarify patients’ preferences and improving surrogate confidence and accuracy in decision-making, thereby reducing stress, decisional conflict and emotional burden.(4, 5) Dyadic engagement has also been linked with improved patient and family outcomes such as improved quality of life near death, greater satisfaction with care, and better bereavement adjustment among family members.(6–8) EOL care decisions are complex and emotionally charged, requiring balancing patient and family’ values and preferences against uncertain prognoses unsupported caregivers experience significant decisional regret and uncertainty. Poor EOL care is associated with greater likelihood of opting for futile and burdensome intervention.(9–11)

In addition to making EOL healthcare decisions, caregivers are increasingly assuming other decision-making responsibilities toward the later stages of the caregiving trajectory due to patients’ cognitive and/or functional decline.(12) Caregivers are expected to act on behalf of care recipients in key domains such as general welfare (e.g. serving as care provider), medical (e.g., as a healthcare proxy), and financial decision-making (e.g., as a financial proxy). In Singapore, these responsibilities are conferred through a range of formal delegation arrangements, i.e. through legally binding processes of becoming a donee via Lasting Power of Attorney (LPA), or other delegation mechanisms such as digital authorizations to manage care recipient’s digital accounts. Caregivers use of personal health portals and proxy access systems is increasing,(13) and national digital platforms such as Singapore’s HealthHub provide a “proxy portal” enabling caregivers to manage appointments, order and pay for medications, view test results, and coordinate care. The expansion of caregivers’ proxy access to health records illustrates a shift toward normalizing proxy involvement beyond patients’ instrumental care to broader care decisions. For example, proxies may use the portal to arrange home care services on behalf of care recipients, effecting where and how care should be provided. These developments suggest an underexplored opportunity to leverage digital delegation frameworks to strengthen ACP engagement by familiarizing caregivers with decision-support roles earlier and through a more gentle and structured approach, alongside with access to patient information and more targeted support for facilitating value conversations with their care recipients.

Despite robust literature on the benefits of enhanced dyadic ACP engagement, promoting caregivers’ engagement remains comparatively understudied as compared to that of patients.(14, 15) This gap is particularly concerning in Singapore and other Asian contexts, where EOL decision-making tends to be relational and family-centric rather than autonomy-centric (16, 17) and discussions about death are deemed a cultural taboo and avoided.(18, 19)

We addressed these gaps by first examining patterns of ACP engagement and formal proxy behaviors among caregivers in Singapore. We then analyzed how formal proxy roles varied in relation to ACP engagement as caregivers prepared for surrogate decision-making. We hypothesized that legal proxies will be less strongly associated with ACP engagement as compared to medical digital proxy because the legal proxy role is more distal in terms of healthcare involvement and decision making as compared to the more immediate and tangible responsibilities associated with the medical digital proxy. Lastly, we identify areas of ACP engagement where digital proxies report less uptake.

## Results

### Demographic characteristics summary

A total of 311 participants [mean age 41.9 (±13.7), female (46.9%), Chinese (78.5%), Malay (12.2%) and Indian (4.8%)] completed the survey between December 2022 and February 2023. More than half of the participants were married (56.9%), had a bachelor’s degree or above (58.8%) and most were employed (85.9%) and living in 3-to-5-room public flats (78.1%) (**Appendix Table S2**). The overall participant sample was comparable to the Singapore general population in most key characteristics, except for higher proportion of higher education (58.8% vs 36%) and employment (85.9% vs 66%). Most participants reported assisting another adult with at least one IADL (87.8%), and the demographic characteristics of this group is also presented in **Appendix Table S2**.

Formal proxy behaviors, particularly digital proxies, were strongly associated with engagement in ACP. In our study, instrumental caregivers who took on formal proxy roles were substantially more likely to report at least one SDM ACP engagement behavior – 85.2% among caregivers with any formal proxy roles versus 47.1% among caregivers without any formal proxy roles. Nevertheless, engagement in SDM is fragmented. In the following segment, we expand on the composition of formal proxy behaviors, legal and digital (financial and medical), and their relationship to SDM engagement.

### Patterns of formal proxy behaviors

Overall, close to one-third (29.3%) of caregivers had a legal proxy role via LPA arrangement and 40.7% had taken on formal digital proxy roles in either financial or medical aspects (**Table 1)**. There were significant overlaps between digital delegation roles (**Appendix Table S3**), with 70% (56 out of 80) of formal *medical* digital proxies also taking on *financial* digital proxy roles.

**Table 1.**
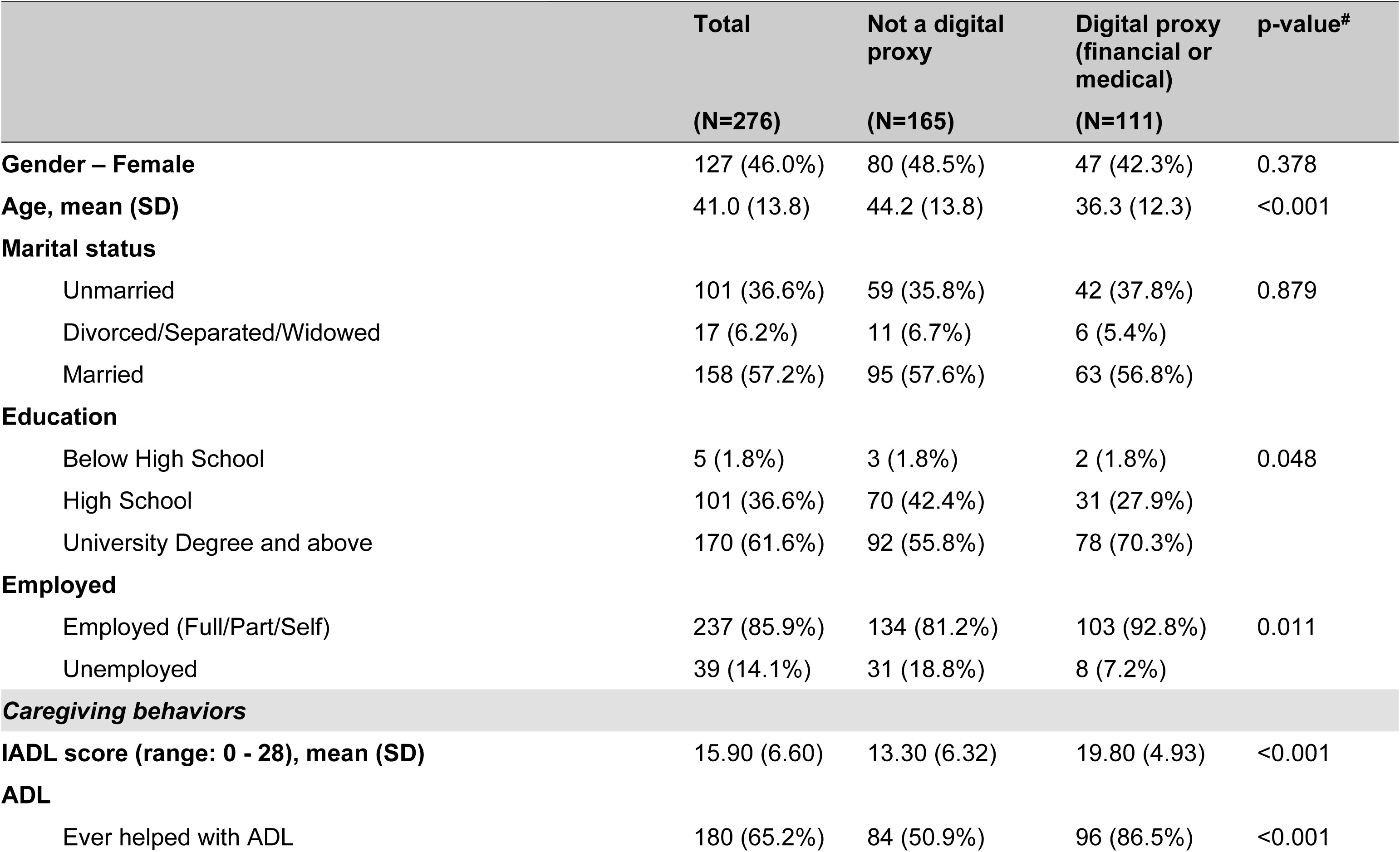
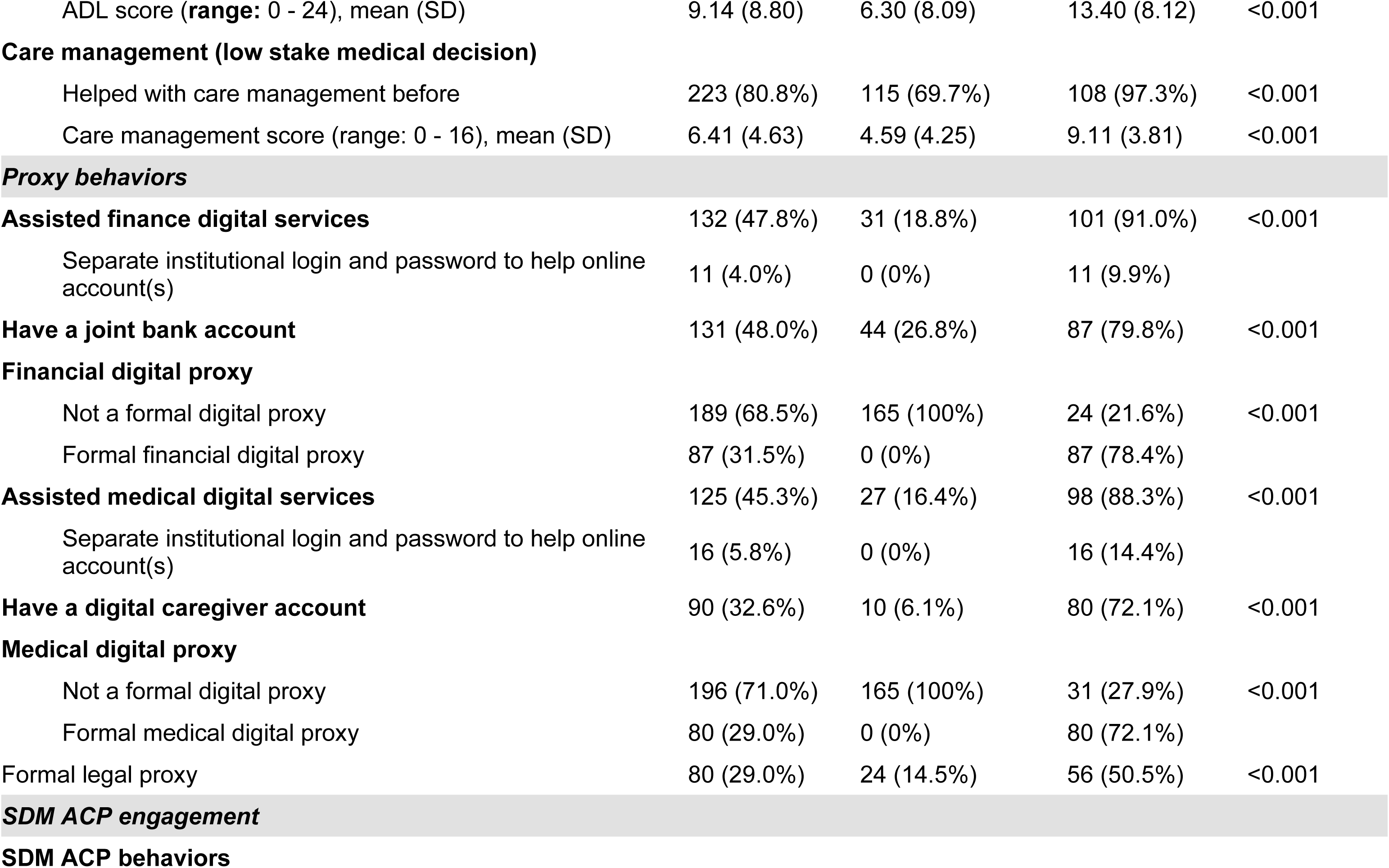
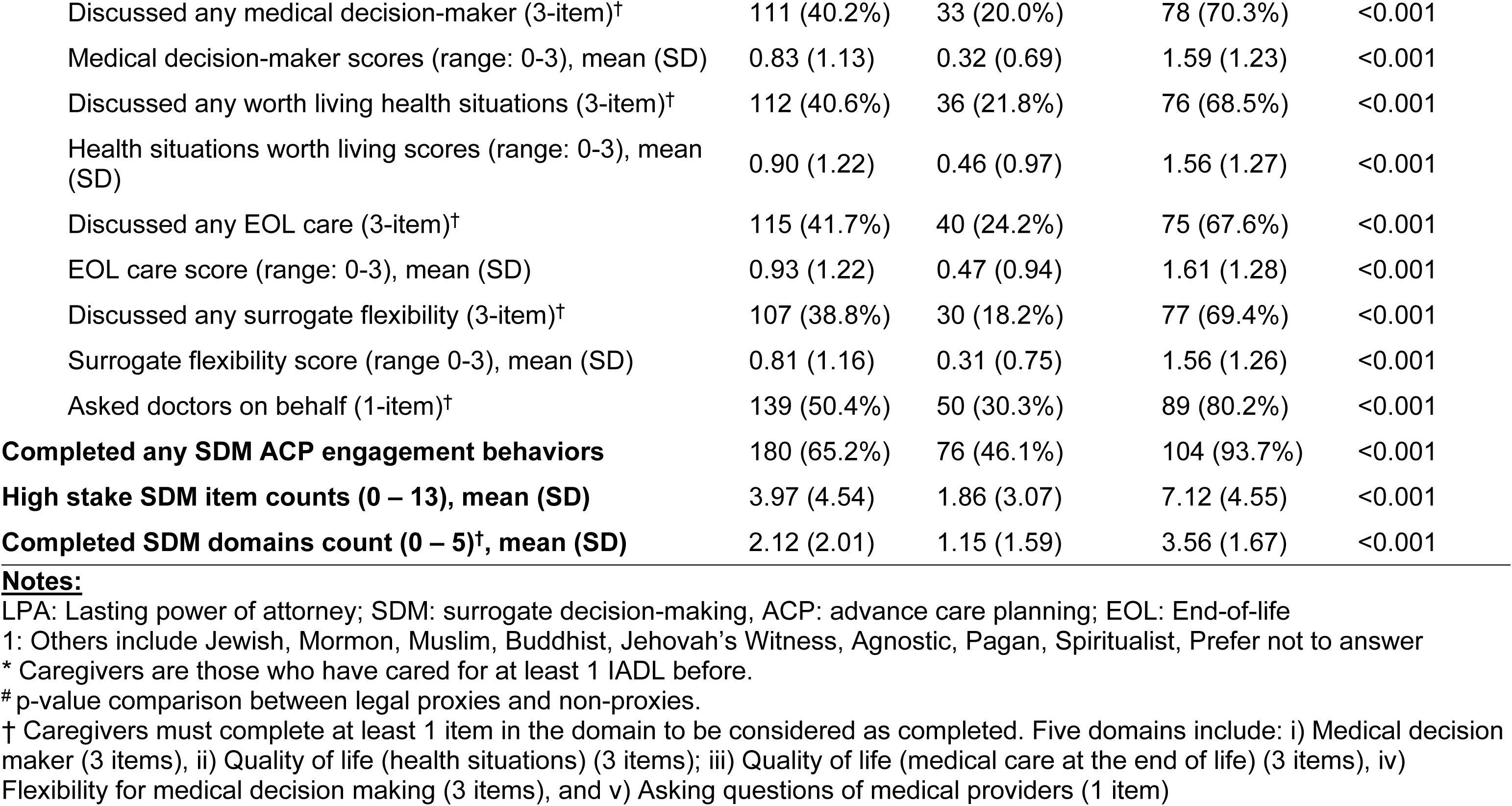
Summary statistics of caregivers.

Among formal digital proxies (n = 111), 27.9% (n = 31) were financial digital proxies only, 21.6% (n = 24) were medical digital proxies only, and 50.5% (n = 56) were both. Among formal financial digital proxies (n = 87), most helped by using a joint bank account only (87.4%, n = 76), while much fewer had authorized access to their care recipient’s digital financial accounts (5.7%, n = 5) or both (6.9%, n = 6).

Among formal medical digital proxies (n = 80), most helped by using a caregiver account only (80.0%, n = 64), while much fewer had authorized access to the care recipients account only (6.2%, n = 5) or had both a caregiver account and authorized access (13.8%, n = 11).

### Strong Relationship between Caregiver Engagement in ACP and Formal Proxy Behaviors

Among caregivers, engagement in ACP was widespread but fragmented, and closely tied to formalized proxy roles. While close to two-thirds (65.2%) reported having completed at least one of the ACP engagement items before, the average number of reported ACP engagement behaviors remained low (3.97±4.54 out of 13 behaviors measured) (**Table 1**). Compared to non-proxy caregivers, caregivers who were digital proxies were more likely to report at least one ACP-engagement behavior (93.7% vs 46.1%, p<0.01), engaged in more SDM items (7.12 vs 1.86, p<0.01), and across more domains (3.56 vs 1.15, p<0.01) (**Table 1** and univariable models in **Table 2**). This pattern also holds when comparing between non-proxies and formal caregivers in the financial and medical domains individually (univariable models in **Table 2**, **Appendix Table S4** and **S5**), and between legal proxies (univariable models in **Table 2**, **Appendix Table S6)**.

**Table 2.**
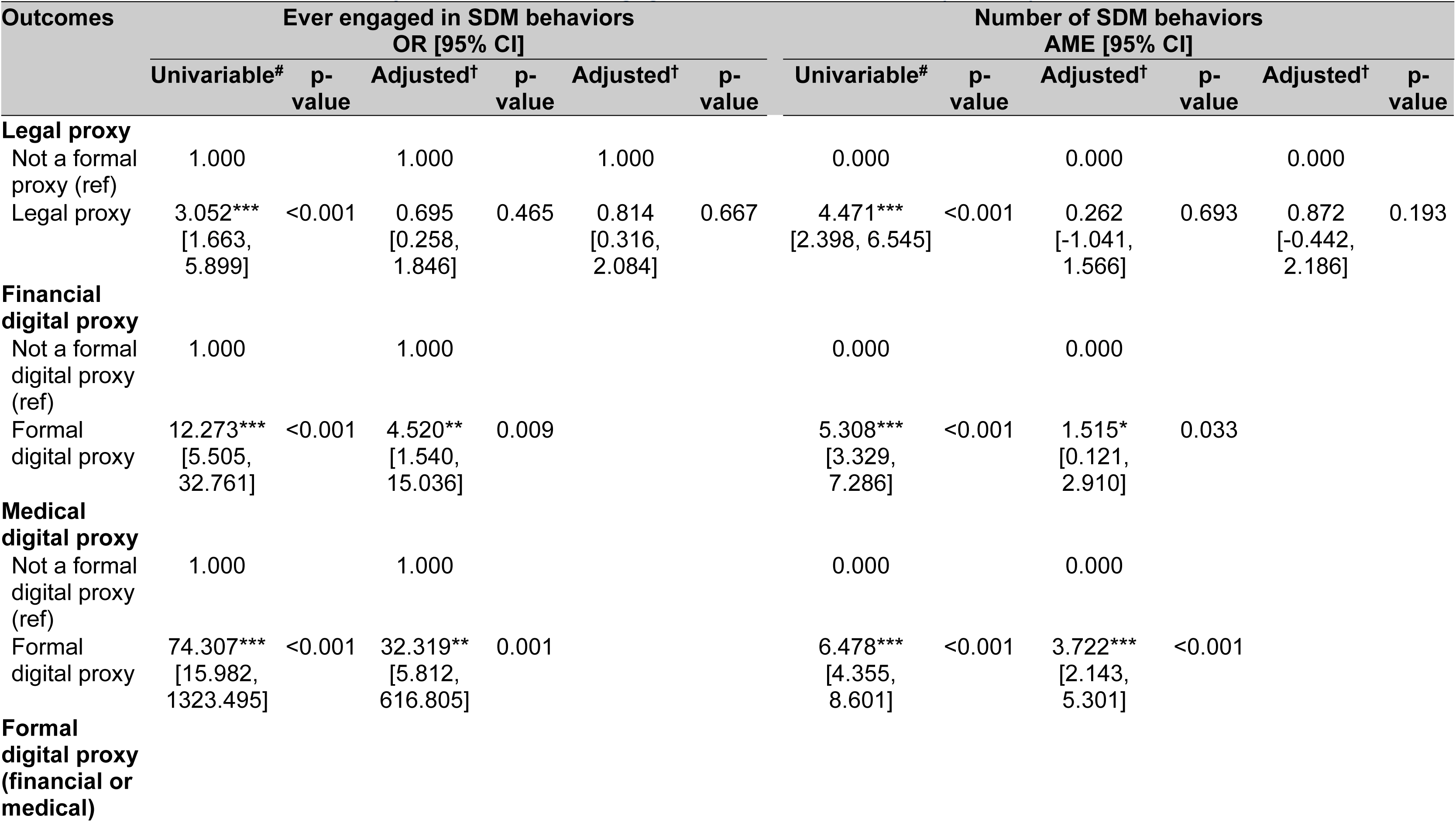
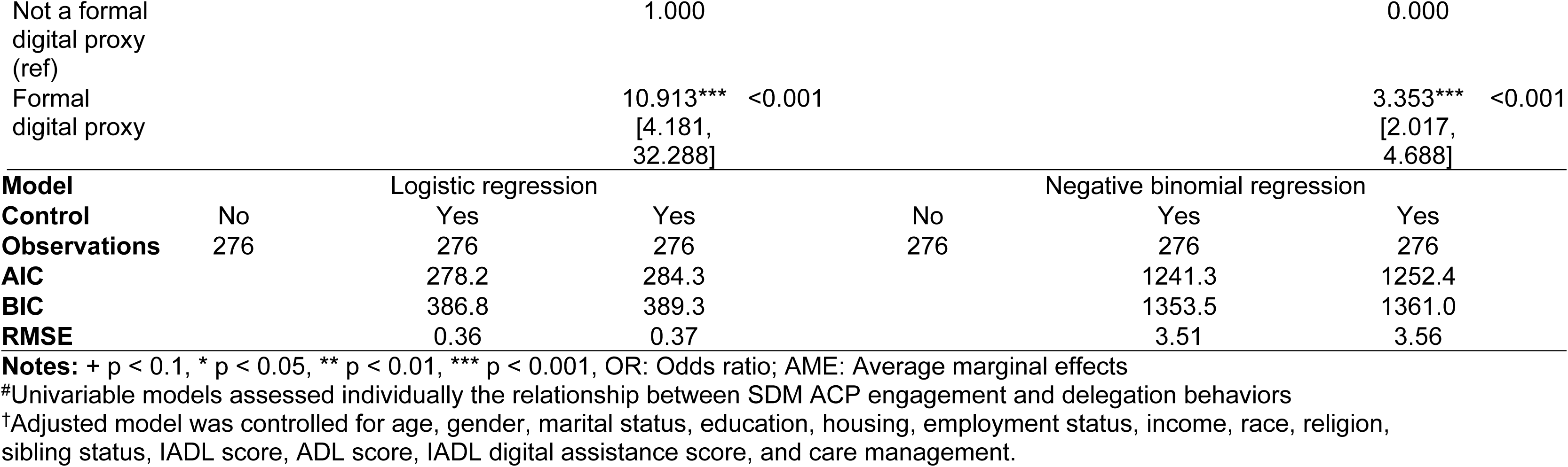
Associations between proxy status and SDM engagement in ACP behaviors. (N = 276)

When adjusted for covariates, medical (OR: 32.319, 95%CI: 5.812 – 616.805, p<0.001) and financial proxies (OR: 4.520, 95%CI: 1.540 – 15.036, p<0.001) were significantly more likely to report ACP SDM behaviors than non digital proxies. Formal medical (AME: 3.722, 95%CI: 2.143 – 5.301, p<0.001) and financial (AME: 1.515, 95%CI: 0.121 – 2.910, p=0.033) digital proxy roles also reported greater extent of SDM ACP engagement as compared to all non-formal digital proxies (**Table 2**). We did not find formal legal proxy arrangement to be associated with greater SDM ACP engagement (OR: 0.695, 95%CI: 0.258 – 1.846, p=0.465; AME: 0.262, 95%CI: -1.041 – 1.566, p=0.693) (**Table 2**).

### Fragmented caregiver ACP engagement

Across proxy roles, we observed a graded pattern of ACP engagement across proxy types (**Figure 1**). Caregivers who were both legal and digital proxies reported the highest engagement across all domains (average 86.4%), followed by those who were formal digital proxies only (average 55.6%), those who were formal legal proxies only (average 27.5%), and lastly non-proxies (average 22.6%). Across all proxy categories except for legal proxy only, asking medical questions on behalf of the care recipients was the most reported SDM ACP behaviors compared to other subdomains.

**Figure 1.**
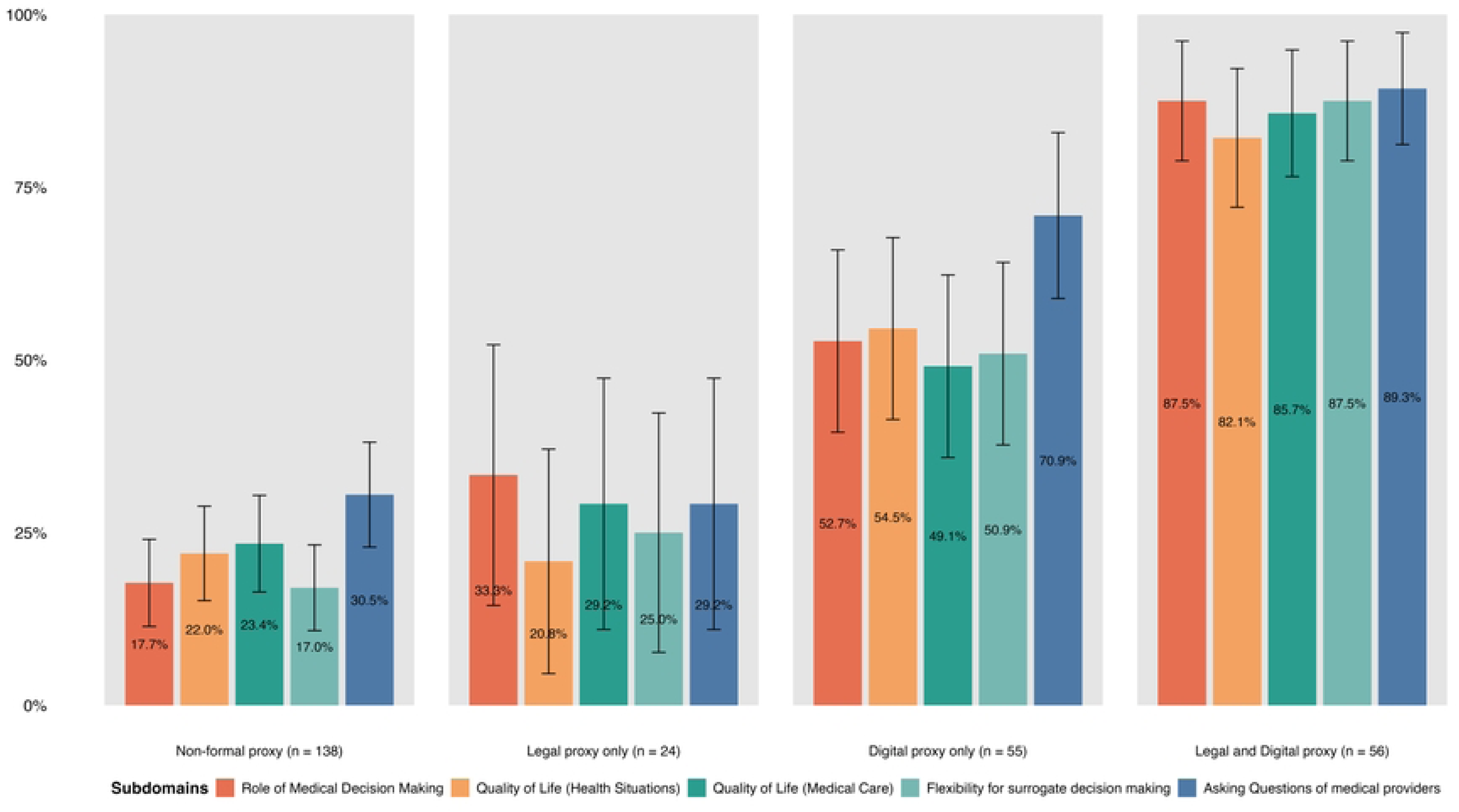
ACP engagement by subdomains completion.

Among non-formal proxy caregivers, the lowest engagement was reported in discussing the flexibility of surrogate decision making (17.4%), followed by the role of surrogate medical decision making (18.1%). Among legal proxies only, the lowest engagement was reported for discussion surrounding what constitutes a life worth living with the care recipient, other family members, or the care team (20.8%) while the highest was the discussion about the role of surrogate medical decision making (33.3%). Among digital proxies only, lowest engagement was reported for discussions surrounding the type of medical care for care recipients (49.1%). Last, similar to legal proxies only, caregivers who were both legal and digital proxies reported the lowest engagement in discussing the health situations considered as life worth living with the care recipient, other family members, or the care team (82.1%).

## Discussion

While previous research shows that greater dyadic engagement in ACP helps to provide value-concordant care,(1) while reducing caregiver decisional conflict and psychological distress,(5, 19) this study found that overall engagement in SDM ACP remained low. In our study, although nearly two-thirds of caregivers reported completing at least one of the ACP engagement behaviors before, the average engagement remained low at 4 out of 13 behaviors, indicating limited depth of ACP engagement. Previous research by Phua et al. has shown a higher-than-expected prevalence of adults in Singapore reported being involved in a high-stake medical ACP engagement task before.(20) However, to our knowledge, this is the first study to quantify the *extent of ACP engagement* specifically among caregivers in Singapore. In the broader context, there have been national initiatives over the past decade, such as public education, hospital-based facilitation and community outreach to promote public engagement in ACP. These efforts have resulted in modest improvements in ACP documentation, but the process continues to face low population-level awareness and engagement.(21, 22) Our findings underscore these engagement gaps.

On the other hand, we found strong positive correlations between formal proxy behaviors, particularly digital proxy, and ACP engagement. We further found that medical digital proxy behavior was the strongest predictor of ACP engagement, and significantly stronger than legal proxy behavior. This suggests that the legal instrument of the LPA may function as a more distal proxy nudge for broader preparatory behavior rather than a direct signal of ACP discussions. For example, the standard LPA Form 1 in Singapore allows for the appointment of personal welfare donees, but it provides limited guidance on how decisions should be grounded in patients’ values. Furthermore, donees’ are restricted in their EOL care decision capacity as they cannot make decisions regarding life-sustaining treatment or treatments that may be aggressive but are needed to prevent serious deterioration of the patient’s health, where advance medical directives and physicians retain ultimate clinical authority.(23) In practice, doctors often still seek the informal consensus of family for EOL decisions.(24) These complexities leave donee caregivers confused about the legal framework for healthcare decisions and even feeling *disempowered*, potentially dampening their motivation to engage in ACP if they believe their input might be overridden at the critical moment.(25) As such, although LPAs may still act as a bridge to ACP by highlighting to donees to consider the donor’s prior wishes, values and preferences, this study suggest that they function more as legal safeguards than as active prompts for values-based conversations.

In contrast, medical digital proxies are embedded within healthcare systems and may serve as a more natural and practical trigger for care and value discussions. Our results suggest that digital proxy behaviors may serve as strategic entry points for interventions to enhance ACP engagement by leveraging existing patterns of caregiver involvement. As medical digital proxy roles are operationally tied to medical information access and healthcare transactions, these responsibilities may effectively prompt caregivers to reflect more concretely on their responsibilities. As such, these findings have potential policy implications. Encouraging caregivers who have executed an LPA to also formalize healthcare-related digital proxies, or vice versa, may create an additional touchpoint within the healthcare system that can be leveraged to support ACP facilitation. Furthermore, healthcare institutions could identify designated medical digital proxies and provide targeted and timely ACP education, nudges or structured discussion tools to promote ongoing ACP engagement.

Last, we also identified proxy-specific domains of engagement where proxies may experience greater difficulties initiating. Unsurprisingly, discussions regarding “health situations that are worth living” had the lowest completion rates among legal proxies or those who were both legal and digital proxies. This is consistent with literature indicating that this domain is the most directly tied to death-related considerations, which are particularly culturally sensitive and socially taboo in Singapore’s Asian context.(26) On the other hand, non-proxies and digital proxies reported the lowest engagement in discussing the flexibility given for surrogate decision-making relative to other subdomains. These findings suggest that while initial ACP steps are being taken, there is a clear deficit when reaching high-stakes clinical scenarios. This gap highlights a critical need to move beyond promoting general awareness and instead provide targeted tools to support caregivers in navigating high-stakes moral and medical dilemmas. Consequently, while broad interventions are necessary to improve engagement across all subdomains, future policy and clinical frameworks must be specifically tailored to address the unique limitations and decisional thresholds associated with different proxy roles.

### Limitations

This study has several limitations. First, participants were recruited through an online survey panel, which may introduce selection bias toward individuals with higher digital literacy, despite efforts to achieve demographic comparability. In addition, the survey was administered exclusively in English, potentially skewing the sample toward English-speaking, more educated and younger respondents, thereby limiting the generalizability of the findings. Although demographic quotas and constraints were applied during recruitment to approximate population characteristics, the final sample remained somewhat younger, more highly educated, and higher-income than the general population.

Second, data quality concerns are inherent in self-reported measures, particularly as all key behaviors in this study were self-reported. Respondents, especially caregivers, may have over-reported shared decision-making (SDM) engagement due to recall bias, recency bias and to a lesser extent, social desirability bias.(27) Because participants were asked about lifetime (“ever”) experiences, those with more recent caregiving roles may have reported behaviors more accurately than those whose caregiving occurred further in the past (28). This weakness is countered by the saliency effects increasing the reporting of key events. Moreover, both LPA and digital proxy arrangements are relatively recent developments. Caregivers in earlier periods may not have had exposure to these mechanisms, which could influence reported prevalence and engagement patterns and bias the association results away from the null hypothesis. Publicly available estimates suggest that approximately 14% of older adults have an LPA, substantially lower than the 30% observed in our sample; however, these figures may not be directly comparable, as our estimate reflects donee status, whereas publicly reported figures refer to prevalence of donors (29).

Finally, we did not assess the regularity or quality of ACP conversations and engagement. Both dimensions are critical, as patients’ and caregivers’ values and preferences are dynamic rather than static, necessitating ongoing, high-quality conversations rather than one-time discussions.(30–32) Emerging evidence further indicates that the depth and quality of values-based discussions may be more consequential than the mere occurrence of a conversation.(33, 34) Future research should therefore examine whether regular review of formal delegation arrangements can facilitate sustained ACP engagement and enhance the quality of these discussions over time.

### Conclusion

As population ageing accelerates and the prevalence of cognitive impairment rises, formal digital delegation represents a promising avenue for enhancing ACP engagement among caregivers. Healthcare systems should consider integrating digital medical delegation mechanisms alongside existing legal frameworks for proxy decision-making. In addition, timely, tailored, and targeted interventions should be directed at these digitally designated proxies to ensure they are adequately prepared to engage in values-based discussions and navigate complex and emotionally challenging EOL care decisions.

## Materials and Methods

### Study design

We fielded a cross-sectional survey to an online panel of resident adults in Singapore. The online panel was created and managed by Qualtrics who recruited panel members from the general population. Panel members were invited to participate in study surveys via email which indicated the study description, research purpose, incentives and duration, or through advertisements on the respondents’ login panel. The inclusion criteria were age 21 years old and above, and English-speaking. To minimize self-selection bias, the survey invitations were kept general without specific details about the survey. We also imposed demographic constraints and quotas (i.e. age groups and gender) during recruitment in efforts to ensure that the respondent sample was similar to the general population.

The survey questionnaire included demographic questions relevant to digital finance and medical proxy roles, and surrogate decision-making (SDM) ACP engagement. Demographic questions included age, gender, education, religion, income, housing, siblings, birth order, employment, marital status and ethnicity. To minimize missing responses, completeness checks were built into the survey instrument. Backward navigation was allowed and respondents could review and change their responses at any point during the survey. The full survey is available in **Supplementary Material Survey Instrument**.

The study was reviewed by the institutional review board of the National University of Singapore with approval number NUS-IRB-2022-412. Respondents indicated consent via a web-based form prior to the survey. Respondents were compensated for completing surveys with selectable rewards including cash, vouchers or coupons by the survey company.

### Formal proxy delegation behaviors

We identified formal proxy roles in three areas: legal proxies, medical, and financial digital proxies. We identified formal legal proxies by asking respondents if they had a signed LPA with the care recipient. Prior to this question, we explained the definition of LPA and the responsibilities of a donee. Those with a signed LPA as a donee were classified as a formal legal proxy. Formal digital proxies are caregivers who have formal access to the care recipient’s digital accounts and are authorized to perform specific actions on behalf of the care recipient. Financial digital proxies are those who reported having “assisted another adult with their legal/financial digital services” *and* either had at least one joint bank account with the care recipient or had institutional access via a separate account to help manage the care recipient’s online accounts (**Table 1**). On the other hand, medical digital proxies are those who reported having “assisted another adult with their digital medical services”, *and* either had a digital caregiver account (that granted access to care recipient’s medical records) or were given institutional access via a separate account (**Table 1**).

### Engagement in ACP

We measured caregivers’ engagement in ACP using items adapted from the 15-item ACP Engagement instrument developed by Sudore et al.(35) The final thirteen items used in the survey instrument included thirteen items comprised the “Action” items from the original instrument and spanned across all five domains of ACP engagement including: i) Medical decision maker (3 items), ii) Quality of life (health situations) (3 items); iii) Quality of life (medical care at the end of life) (3 items), iv) Flexibility for medical decision making (3 items), and v) Asking questions of medical providers (1 item) (**Appendix Table S1**). Response options included ‘Yes,’ ‘No, but I should have,’ and ‘No, because I didn’t need to’, with the latter two options included to differentiate missed opportunities from the lack of opportunities. Engagement was measured as the total number of items completed, ranging from 0 to 13, with higher score indicating greater engagement. We also examined engagement on domain-level by assessing domains with at least one completed engagement behaviors.

Questions were phrased to ask participants about *any* experience of caregiving responsibilities and ACP engagement behaviors as opposed to recent or current experiences. This allowed us to capture a broader range of caregiving experiences, especially among those with past or episodic caregiving experience. Furthermore, ACP engagement was phrased as “medical delegation action (in high-stake decisions)” to avoid negative connotations often associated with ACP.(36)

### Caregiving intensity

We measured caregiving intensity as indicators of the functional health/wellbeing of the care recipients and extent of caregiving involvement. We adapted the Instrumental Activities of Daily Living (IADL) scale,(37) and the Activities of Daily Living (ADL) scale.(38) We also included an item on assisting with digital tasks (such as logging into mobile apps, operating applications, checking emails and changing device settings) to measure participants’ daily digital functioning given the increasingly digital living environment. Participants were asked to report the highest frequency of help in both physical and digital tasks they had provided to the care recipients in each activity, ranging from daily (0), weekly (1), monthly (2), yearly (3), to never (4). Total scores were calculated (ranging from 0 to 28) where a higher total score indicated greater intensity of caregiving. We also measured the intensity of care-management activities, including helping make appointments, accompanying the care recipient to medical appointments, ensuring proper medication intake and medical treatment procedures. Similar to ADL and IADL items, participants were asked to describe the highest frequency they have assisted in such tasks, ranging from never (0) to daily (4). Total score was calculated, ranging from 0 – 16 where a higher score indicated greater intensity of care-management.

### Statistical analysis

We first summarized the characteristics of the sample. We assessed the association between formal proxy roles (i.e. LPA status, formal financial and medical digital proxies) and extent of SDM engagement (i.e. number of SDM behaviors reported) using negative binomial regression. Adjusted models were controlled for proxies’ sociodemographic characteristics (age, gender, ethnicity, education, marital status, employment, housing situation, sibling order)(39) and caregiving intensity (ADL, IADL, and care-management activities).(40) We limited our analyses to only participants who reported assisting another adult with at least one IADL (88% of the sample and henceforth termed ‘caregivers’), as such assistance often corresponds with initiation of broader decision-making responsibilities and rights, and potentially, ACP engagement.(41) Last, we identified specific engagement gaps that may benefit from additional support and targeted interventions. All analyses were performed using R version 4.5.0. Model performance and goodness-of-fit were assessed using McFadden’s Pseudo R^2^ and the Akaike Information Criterion (AIC). All statistical tests were two-tailed, and statistical inferences were conducted using a 95% confidence interval, with a significance threshold set at a p-value<0.05. This study is reported in accord with the Strengthening the Reporting of Observational Studies in Epidemiology (STROBE) statement for cross-sectional studies. A completed STROBE checklist for cross-sectional studies is available in the supplementary files (**Appendix STROBE checklist**).

## Author contributions

CZ, PVSP and PSF devised the project and conceptual ideas and designed and implemented the survey. VAH and PSF jointly crafted the research questions. VAH performed the statistical analyses. VAH and PSF drafted the manuscript. CZ, PVSP, and GCHK critically revised the manuscript. GCHK obtained funding for the study. PSF is the guarantor of this research. All authors have agreed on the final version and meet at least one of the following criteria (recommended by the ICMJE: http://www.icmje.org/ethical_ 1author.html).

## Acknowledgement

We thank Natasha Ureyang for their feedback on the manuscript.

## Funding Declaration

This work was supported by the Singapore Ministry of Heath’s National Medical Research Council Grants (grant numbers NMRC/CG1/009/2022-NUH and CareEco21-0030). The funders had no role in study design, data collection and analysis, decision to publish, or preparation of the manuscript.

## Competing interests

The authors declare no competing interests.

## Declaration

### Declaration of generative AI and AI-assisted technologies in the manuscript preparation process

During the preparation of this work the authors used ChatGPT-5.3 to assist with rewording in drafting the manuscript and enhancing the overall readability. After using this tool/service, the authors reviewed and edited the content as needed and take full responsibility for the content of the published article.

### Data availability

All data and relevant information are with the paper and its supplemental files.

### Code availability

All the statistical analyses were conducted using R version 4.5.0. Codes are not publicly available but may be made available from the corresponding authors on reasonable request.

## Appendix title and captions

**Appendix Table S1. Surrogate decision-maker engagement in advance care planning measures**

**Appendix Table S2. Summary of demographics of survey respondents and comparison to general population, stratified by caregiving status**

**Appendix Table S3. Distribution of digital delegations**

**Appendix Table S4. SDM ACP engagement by formal medical digital proxy status**

**Appendix Table S5. SDM ACP engagement by formal financial digital proxy status**

**Appendix Table S6. SDM ACP engagement by legal proxy status**

## Notes

### Competing Interest Statement

The authors have declared no competing interest.

### Author Declarations

The study was reviewed by the institutional review board of the National University of Singapore with approval number NUS-IRB-2022-412.

